# Higher contact among vaccinated can be a mechanism for negative vaccine effectiveness

**DOI:** 10.1101/2022.04.25.22274266

**Authors:** Korryn Bodner, Jesse Knight, Mackenzie A. Hamilton, Sharmistha Mishra

## Abstract

Evidence from early observational studies suggested negative vaccine effectiveness for the SARS-CoV-2 Omicron variant. Using transmission modeling, we illustrated how increased contact between vaccinated individuals, vaccinated contact heterogeneity, paired with lower vaccine efficacies could produce negative measurements and how we can identify this mechanism via a key temporal signature.

## Introduction

Within 4 weeks of the emergence and in the context of rising cases of Omicron, population-based studies in Canada [1], Denmark [2], and the United Kingdom [3] had reported “negative vaccine effectiveness” against SARS-CoV-2. Vaccine effectiveness (V_Eff_) is calculated by comparing the rates of infection between vaccinated and unvaccinated individuals. Thus, a negative V_Eff_ measurement suggests that vaccinated individuals were acquiring infections at higher rates than unvaccinated individuals. One potential explanation for the increased infection was that the vaccine increased biological susceptibility, for example, if the virus had evolved to spread faster in vaccinated individuals [4]. However, V_Eff_ measurements are calculated using observational data and thus subject to various biases, including but not limited to differences in testing/detection and exposures among vaccinated and unvaccinated populations [5]. Differential exposures by vaccination status could stem from contact heterogeneity.

Contact heterogeneity refers to different levels of contact among and between population subgroups. Increased contact between vaccinated persons, potentially arising due to policies that restrict certain spaces to vaccinated individuals (e.g. vaccine mandates), is one type of contact heterogeneity (hereafter, vaccinated contact heterogeneity). In this study, we test (1) whether vaccinated contact heterogeneity could lead to negative V_Eff_ measurements; (2) how this relationship is affected by two components of vaccine efficacy related to transmissibility: vaccine efficacy against susceptibility (VE_S_) and vaccine efficacy against infectiousness (VE_I_) [6]; and if negative measurements can be produced, (3) how this mechanism can be identified. VE_S_ and VE_I_ reflect the true total vaccine benefit against infection, with VE_S_ reflecting the reduced probability of vaccinated recipients acquiring infection and VE_I_ reflecting the reduced infectiousness of vaccinated individuals if a breakthrough infection occurs. We hypothesize that both vaccinated contact heterogeneity and the levels of VE_S_ and VE_I_ contribute to producing negative V_Eff_.

## Methods

Following Shim and Galvani [7], we adapted a simple compartmental SIR (susceptible, infectious, recovered) transmission dynamics model for vaccinated and unvaccinated individuals that assumed an all-or-nothing vaccine type (supplementary Figure 1, [8]). To explicitly account for potential contact differences, the SIR model contained both within-group contact rates for unvaccinated, *c*_*uu*_, and vaccinated individuals, *c*_*vv*_, as well as between-group contact rates for unvaccinated with vaccinated, *c*_*uv*_, and vaccinated with unvaccinated, *c*_*vu*_.

In all simulations, we assumed 75% vaccination coverage. We explored two different contact scenarios: *homogeneous contact*, where vaccinated and unvaccinated individuals have equal contacts with random (“proportionate”) mixing; and *vaccinated heterogeneous contact* where vaccinated individuals have increased within-group contact. In the *homogenous contact* scenario, we assumed 6 daily contacts per-person, reflecting approximate contact rates from U.S. and U.K. during the pandemic [9], and thus defined *c*_*vv*_ = *c*_*uv*_ = 4.5 and *c*_*uu*_= *c*_*vu*_=1.5. In the *vaccinated heterogenous contact* scenario, contacts between vaccinated were increased by 50% compared to the *homogeneous contact* scenario (*c*_*vv*_ = 6.75), with all other parameter values unchanged. We set the recovery rate to be 1/10 [8] and the probability of transmission to be 0.01 such that R_0_ = 6 in a fully unvaccinated population with random mixing. Given the uncertainty surrounding vaccine efficacies, two different baseline values of VE_I_ and VE_S_ were adopted (0.1, 0.5). We also conducted sensitivity analyses, varying VE_I_ and VE_S_ from 0.1 to 1 and increasing *c*_*vv*_ by 0%-100% from the *homogenous contact* scenario rates (*c*_*vv*_ = 4.5 - 9). To start our simulations, we introduced one infected vaccinated and unvaccinated individual into our population.

Following Haber [9], we measured V_Eff_(t) as 1 – relative risk (t) (RR[t]), defined as:

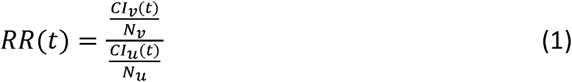

where *Cl*_*v*_ *(t)* and *Cl*_*u*_*(t)* are the cumulative incidences for vaccinated and unvaccinated groups at time *t* and *N*_*v*_ and *N*_*u*_ are the total numbers of vaccinated and unvaccinated individuals, respectively. We also tracked how differences in the depletion of the proportion of susceptible vaccinated, 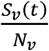, and unvaccinated, 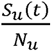, interacted explicitly with VE_S_ to influence measurements of V_Eff_(t) (supplementary Material 1). R-code to recreate all analyses is available on GitHub (https://github.com/kbbodner/contact_and_NegVE)

## Results

First, scenarios of homogeneous contact by vaccination status never led to an observed negative V_Eff_. Second, scenarios of heterogeneous contact by vaccination status produced negative V_Eff_, but only in the context of lower vaccine efficacies (VE_S_ =0.1, VE_I_=0.1, and VE_S_=0.1, VE_I_=0.5; Figure 1a). Third, negative V_Eff_ only occurred during epidemic growth (Figure 1a and b) with V_Eff_(t) becoming positive only when the proportion of susceptible unvaccinated was lower than the combined proportion of susceptible vaccinated with the proportion immune due to vaccination (i.e. the level of VE_S_; supplementary Figure 2).

**Figure 1.**
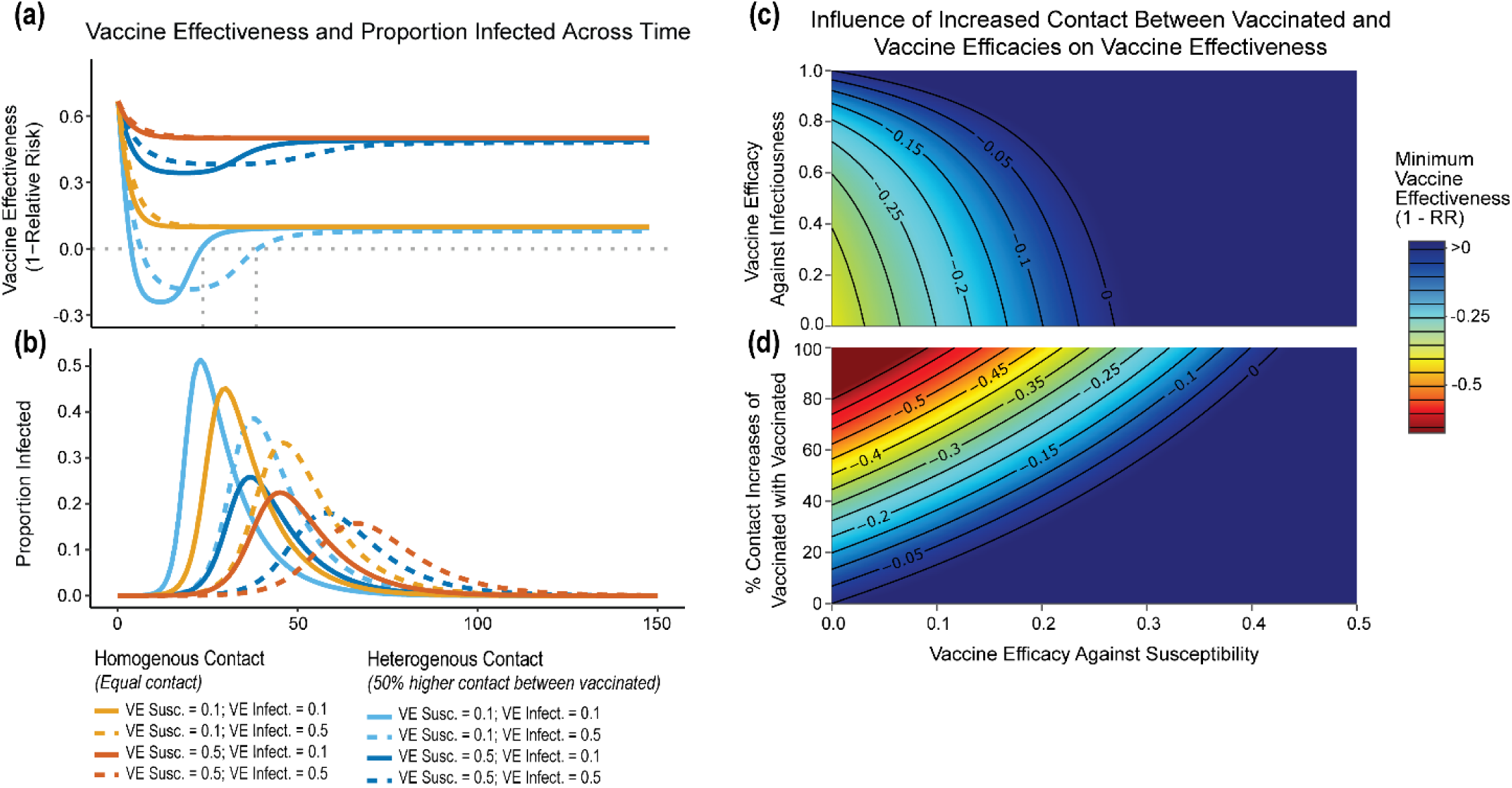
Vaccine effectiveness and infection dynamics are influenced by contact heterogeneity and vaccine efficacies. Homogeneous contact rates (equal contacts among vaccinated and unvaccinated individuals) and heterogenous contact rates (vaccinated have more contacts with vaccinated individuals) interact with vaccine efficacy against susceptibility (VE_S_) and vaccine efficacy against infectiousness (VE_I_) to influence measurements of vaccine effectiveness over time (a) and the proportion of infected individuals over time (b). Negative vaccine effectiveness becomes positive once the proportion of susceptible unvaccinated individuals became lower than the proportion of susceptible vaccinated individuals combined with the level of VE_S_ (grey vertical lines; supplementary Material 1). The minimum vaccine effectiveness was sensitive to VE_I_ (c), the % increase in contact between vaccinated individuals (d), and VE_S_ (c and d). Note that colours in (c) and (d) indicate the minimum negative vaccine effectiveness observed for a given simulation with >0 indicating a non-negative measurement.

The minimum V_Eff_ was moderately influenced by VE_I_, and strongly influenced by the levels of VE_S_ and the contact between vaccinated individuals (Figure 1c-d). For example, when VE_S_ was less than 0.2 and *C_vv_* was a 100% higher than the *homogeneous contact* scenario, V_Eff_(t) was strongly negative (< - 0.5). VE_I_ was less influential on negative V_Eff_ but high levels of VE_I_, (>0.92) could still prevent negative V_Eff_ even at very low VE_S_ (<0.1) (Figure 1c).

## Discussion

Our results demonstrated how vaccinated contact heterogeneity, defined as higher contact levels between vaccinated individuals, could lead to observed measurements of negative V_Eff_. Thus, we illustrate a plausible scenario where vaccines can be perceived to be non-beneficial - or even harmful – despite providing a benefit to a population (vaccine efficacies > 0).

Vaccinated contact heterogeneity can negatively bias measurements of V_Eff_, but observing negative measurements required the underlying vaccine efficacies to be lower– in particular, lower VE_S_. That is, we found that vaccine efficacies can mediate the effect of the contact heterogeneity bias. Given the consistent reports of higher V_Eff_ against other variants compared to Omicron [e.g. 10], this mediation effect can explain how this bias could be present before Omicron despite the absence of negative measurements.

Beyond testing vaccinated contact heterogeneity feasibility as a mechanism of bias, we also identified a temporal signature in V_Eff_ measurements that indicates when this mechanism could be the cause of negative V_Eff_. In the context of vaccinated contact heterogeneity, negative measurements only occurred during epidemic growth when the proportion of susceptible unvaccinated was higher than the proportion of susceptible vaccinated (mediated by VE_S_; supplementary Figure 2). In each of the empirical studies, the negative V_Eff_ measurements coincided with Omicron’s epidemic growth stage [1–3]. If measurements of V_Eff_ are consistently updated and found to change direction later in an epidemic, this would suggest the negative measurement may have been the result of vaccinated contact heterogeneity.

Vaccinated contact heterogeneity is one possible cause of negative V_Eff_, but other biases such as selection bias via testing access or health-seeking behaviour [5], as well as higher immunity among unvaccinated from prior infection could also potentially cause negative measurements. Moreover, our analysis focused on an all-or-nothing vaccine type for simplicity, but leaky vaccine type [11] could impart a different temporal pattern for the vaccinated contact heterogeneity bias. Important next steps include exploring other potential biases that may lead to negative V_Eff_ and including how assumptions surrounding leaky versus all-or-nothing vaccine type may influence V_Eff_ measures over time.

Although our study was designed to explain potential mechanisms, and not to specify which values of VE_S_, VE_*I*_ and contact differences most likely cause observed negative measures, the findings have important implications for the conduct and interpretation of observational studies measuring V_Eff_. If possible, observational studies must try to address confounding due to vaccinated contact heterogeneity when measuring V_Eff_ during epidemic growth as this bias could affect measurements for future variants of SARS-CoV-2 and other emerging pathogens. If it is not possible to address confounding, then reports and public communication must ensure that interpretation of V_Eff_ includes the possibility of this bias to avoid misinterpretation that can amplify vaccine mistrust [12], or wait until the epidemic peak has occurred to update and report measurements of V_Eff_.

In this brief report, we highlight one possible pathway for V_Eff_ to appear negative even when vaccines are beneficial and how this bias could be identified. Our findings not only illustrate a potential mechanism for the negative V_Eff_ studies of the Omicron variant [1,3], but also provide a potential explanation for observed negative V_Eff_ in future studies.

## Data Availability

Simulated data for this study are available on GitHub.

https://github.com/kbbodner/contact_and_NegVE

## Funding

This work was supported by the Canada COVID-19 Immunity Task force grant (to SM).

## Acknowledgements

We would like to thank Dr. Jeffrey C. Kwong and Sarah A. Buchan for their insight into measuring vaccine effectiveness and KB would like to additionally thank Cedric B. Hunter for their ongoing support. JK is supported by the Natural Sciences and Engineering Research Council of Canada (NSERC CGS-D) and SM is supported by a Tier 2 Canada Research Chair in Mathematical Modeling and Program Science.

## Notes

This work was supported by the Canada COVID-19 Immunity Task Force grant (to SM) All authors declare that they have no conflicts of interest.

### Competing Interest Statement

The authors have declared no competing interest.

### Funding Statement

This study was funded by the Canada COVID-19 Immunity Task Force grant

### Summary of Updates

Small modification to supplementary material 1 and provision of GitHub repository link to recreate simulations

