## supplementary for "Higher contact among vaccinated can be a mechanism for negative vaccine effectiveness"

**Supplementary Material 1**

**SIR transmission model**

We used an SIR (susceptible, infectious, recovered) compartmental transmission dynamics model to perform our analyses (Figure S1). The model equations (coupled ordinary differential equations) are as follows:

$\frac{ⅆS_{u}}{ⅆt}=-\beta S_{u}\left( c_{uu}\frac{I_{u}}{N_{u}}+\left( 1-\mathrm{VE}_{I} \right)c_{uv}\frac{I_{v}}{N_{v}} \right)$	[1]

$$\frac{ⅆI_{u}}{ⅆt}=\beta S_{u}\left( c_{uu}\frac{I_{u}}{N_{u}}+\left( 1-\mathrm{VE}_{I} \right)c_{uv}\frac{I_{v}}{N_{v}} \right)-I_{u}\gamma$$

$$\frac{ⅆR_{U}}{ⅆt}=I_{u}\gamma$$

$$\frac{ⅆS_{v}}{ⅆt}=-\beta S_{v}\left( c_{vu}\frac{I_{u}}{N_{u}}+\left( 1-\mathrm{VE}_{I} \right)c_{vv}\frac{I_{v}}{N_{v}} \right)$$

$$\frac{ⅆI_{v}}{ⅆt}=\beta S_{v}\left( c_{vu}\frac{I_{u}}{N_{u}}+\left( 1-\mathrm{VE}_{I} \right)c_{vv}\frac{I_{v}}{N_{v}} \right)-I_{v}\gamma$$

$$\frac{ⅆR_{v}}{ⅆt}=I_{v}\gamma$$

where $S_{u}$, $I_{u}$, and $R_{u}$ are the numbers of susceptible, infected, and recovered unvaccinated individuals, respectively; $S_{v},I_{v}$ and $R_{v}$ are the corresponding numbers for vaccinated individuals; $N_{u}={S_{u}+I_{u}+R}_{u}$ and $N_{v}={S_{v}+I_{v}+R}_{v}$, which are the total numbers of unvaccinated and vaccinated individuals respectively; $\beta$ is the probability of infection per contact; $\gamma$ is the recovery rate; $\mathrm{VE}_{I}$ is the vaccine efficacy against infectiousness; $c_{uu}$ and $c_{vv}$ are the within-group contact rates for unvaccinated and vaccinated individuals, respectively, and $c_{uv}$ and $c_{vu}$ are the contact rates of unvaccinated with vaccinated and vaccinated with unvaccinated, respectively.

The initial conditions for vaccinated individuals included vaccine efficacy against susceptibility, $\mathrm{VE}_{S},$and were defined as:

$S_{v}\left( t=0 \right)=\left( 1-\mathrm{VE}_{S} \right)N_{v}-\varepsilon$,

$I_{v}\left( t=0 \right)=\varepsilon,$

$$R_{v}\left( t=0 \right)=\mathrm{VE}_{S}N_{v}$$

Whereas for unvaccinated individuals, our initial conditions were defined as:

$$S_{u}\left( t=0 \right)=N_{u}- \varepsilon,$$

$$I_{u}\left( t=0 \right)=\varepsilon,$$

$R_{u}\left( t=0 \right)=0$.

where $\varepsilon=1$ was the number of infected individuals introduced into each group at $t=0$. The model was coded in R (version 4.1.2; (1)), and solved numerically using the *lsoda* function within the deSolve package (2).

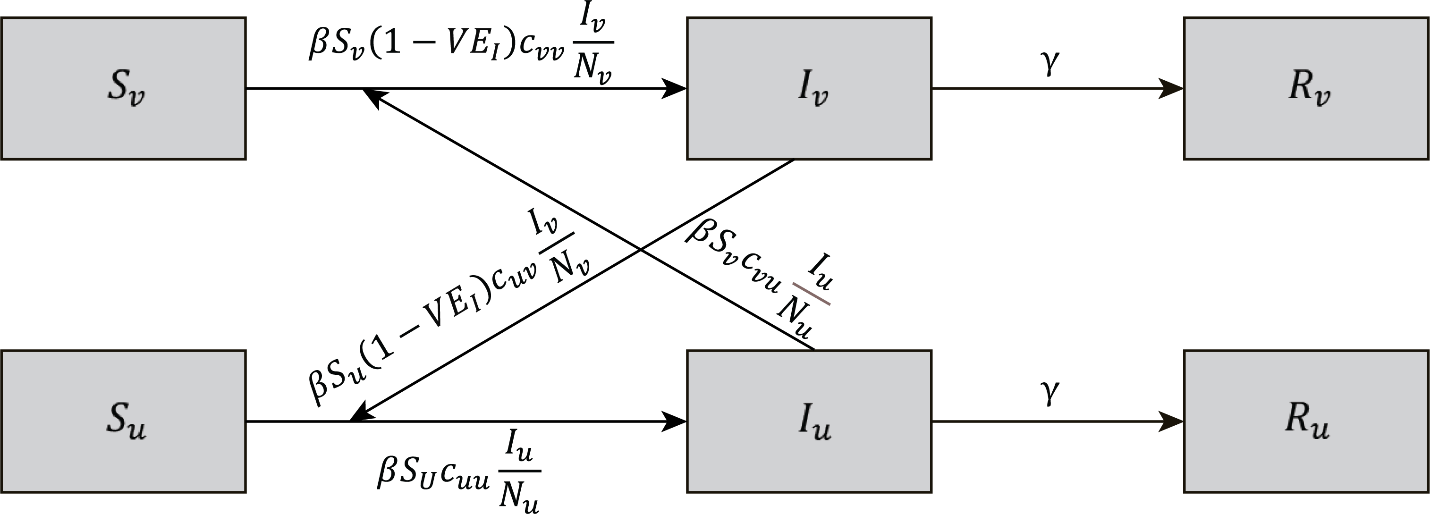

**Figure S1:** A schematic of our SIR compartment model where S, I, and R represent the susceptible, infected, and recovered individuals who are either vaccinated (v) or unvaccinated (u). Susceptible individuals are infected via a transmission probability (β) based on their contact rates with either infected vaccinated (c_uv_ or c_vv_) or infected unvaccinated individuals (c_vu_ or c_uu_). Susceptible vaccinated and unvaccinated individuals have a reduced chance of infection when contacting an infected vaccinated individual due to vaccine efficacy against infectiousness (VE_I_). Protection due to vaccine efficacy against susceptibility (VE_S_) is not included in the main equations but instead is included at the start of the simulation when a specified proportion of vaccinated individuals (defined by VE_S_) is moved to the recovered (and thus immune) compartment. Once infected, all vaccinated and unvaccinated individuals recover from infection at the same rate (γ).

**Derivation of the relationship between vaccine effectiveness, the proportion of susceptible individuals and vaccine efficacy against susceptibility**

Following Haber (3), we define vaccine effectiveness over time, V_Eff_(t) as follows:

$${VE}_{Eff}\left( t \right)=1-RR\left( t \right) [2]$$

with

$$RR\left( t \right)=\frac{\frac{{CI}_{v}\left( t \right)}{N_{v}}}{\frac{{CI}_{u}\left( t \right)}{N_{u}}} [3]$$

where ${CI}_{v}(t)$ and ${CI}_{u}(t)$ are the cumulative incidences for vaccinated and unvaccinated groups at time *t,* respectively. Given equations 2 and 3:

V_Eff_(t) $\geq0$ when $\frac{{CI}_{v}\left( t \right)}{N_{v}}\leq$ $\frac{{CI}_{u}\left( t \right)}{N_{u}}$ $[4]$

and

V_Eff_(t) $<0$ when $\frac{{CI}_{v}\left( t \right)}{N_{v}}$ $>\frac{{CI}_{u}\left( t \right)}{N_{u}}$ $[5]$

We define ${CI}_{v}(t)$ and ${CI}_{u}(t)$ as:

$${CI}_{v}\left( t \right)=I_{v}\left( t \right)+R_{v}\left( t \right) , [6]$$

$${CI}_{U}\left( t \right)=I_{U}\left( t \right)+R_{U}\left( t \right)$$

Let $N_{V}= {S_{v}(t)+ I}_{v}(t)+R_{v}(t)+ VE_{S} N_{v}$ and $N_{U}= {S_{u}(t)+ I}_{U(t)}+R_{u}(t)$ with $N_{v}$ and $N_{u}$ defined as constant over time given no mortality exists in the system. With equations 6 and these definitions, we can rearrange equation 4 as:

$\frac{{N_{V}- S}_{v}\left( t \right)- VE_{S} N_{v}}{N_{v}}\leq$ $\frac{{N_{u}- S}_{u}\left( t \right)}{N_{u}}$	$[7]$

$$1- \frac{S_{v}\left( t \right)}{N_{v}}- VE_{S}\leq1- \frac{S_{u}\left( t \right)}{N_{u}}$$

$$\frac{S_{v}\left( t \right)}{N_{v}}+ VE_{S}\geq\frac{S_{u}\left( t \right)}{N_{u}}$$

Here we find that V_Eff_ becomes non-negative only when the proportion of susceptible unvaccinated, $\frac{S_{u}\left( t \right)}{N_{u}}$, is less than the combined proportion of susceptible vaccinated, $\frac{S_{v}\left( t \right)}{N_{v}}$ , and those vaccinated and immune ($VE_{S}$). Similarly, using equations 6 and the inequality of equation 5, we find that V_Eff_ is negative when $\frac{S_{u}\left( t \right)}{N_{u}}$is greater than the combined proportion of $\frac{S_{v}\left( t \right)}{N_{v}}$ with $VE_{S}$.

In Figure S2, we illustrate that the inequality of equation 4 was not maintained in our vaccinated contact heterogeneity scenarios and that the two inequality changes that occurred in each scenario defined the period of negative V_Eff_ measurements.

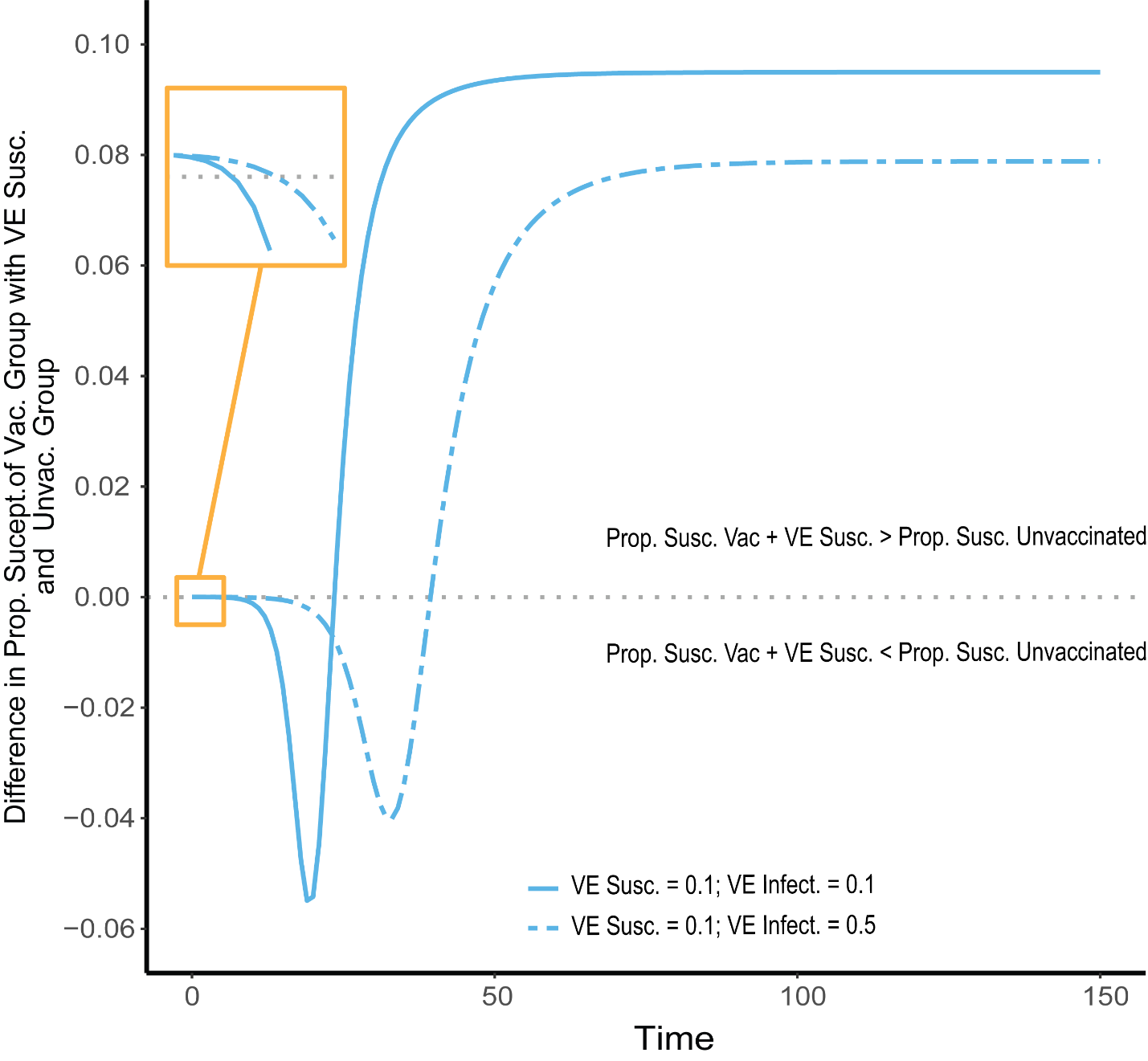

**Figure S2:** A measure of the differences between the proportion of vaccinated susceptibles (Prop. Susc. Vac) and the proportion of unvaccinated susceptibles (Prop. Susc. Unvac.) accounting for vaccine efficacy against susceptibility (VE Susc.) over time. After an initial period, both vaccinated heterogeneous scenarios (VE Susc. =0.1 and VE Infect. =1 [solid line]; VE Susc. =0.1 and VE Infect. = 0.5 [dashed line]) have a higher proportion of susceptible unvaccinated individuals compared to the combined proportion of susceptible vaccinated and immune vaccinated individuals (VE Susc.). This relationship remains until day 25 (VE Susc. =0.1 and VE Infect. =1) and 41 (VE Susc. =0.1 and VE Infect. = 0.5) when this inequality flips with the timing of these crossovers corresponding to when vaccine effectiveness switches from negative to positive (equation 5).
